# The epidemiological investigation of hyperlipidemia in the northern Henan Province

**DOI:** 10.1101/2023.07.24.23293065

**Authors:** Xing Lu, Ziyang Lin, Junzheng Yang

**Affiliations:** Cardiovascular medicine department IV of Xinxiang Central Hospital, Xinxiang Henan 453000, China; Guangdong Nephrotic Drug Engineering Technology Research Center, The R&D Center of Drug for Renal Diseases, Consun Pharmaceutical Group, Dongpeng Avenue, No.71, Guangzhou, Guangdong 510530, China

**Keywords:** hyperlipidemia, epidemiology, prevention and treatment, risk factor, northern Henan Province

## Abstract

**Background/purpose:** Hyperlipidemia is a kind of diseases with the characteristic of high level of blood lipids, it could directly result in several serious diseases including atherosclerosis and coronary heart disease to threat to human health. Therefore, to understand the epidemiological status and risk factors of hyperlipidemia is important for the prevention, diagnosis, and treatment of hyperlipidemia.

**Methods:** The clinical data in Xinxiang Central Hospital from 2019-2021 were collected, and those data were classified and analyzed according to the basic information of investigation subjects and risk factors.

**Results:** A total of 670 investigation subjects were collected in this investigation including 521 males and 149 females, accounting for 77.76% and 22.24% in the total investigation population, respectively; age range of investigation subjects was 20-97 year old; there were 390 people with a history of smoking and 346 people were never smoking, accounting for 58.21% and 51.64% in the total investigation population, respectively; in this investigation, the proportion of peasant farmers was the highest (353/670, 52.69%), followed by retire population (164/670, 24.48%); in terms of education level, the proportion of people graduating from middle school was the highest (338/670, 50.45%), followed by those graduating from primary school (249/670, 37.16%); the statistical results found that the incident of hyperlipidemia were correlated to gender (P<0.01), ethnicity (P<0.01), age (P<0.05), marital status (P<0.05), education level (P<0.001), hypertension (P<0.05) and diabetes mellitus (P<0.001), had no relation to occupation (P>0.05), cerebral hemorrhage, angina pectoris, ischemic stroke, gastrointestinal ulcers, coronary heart disease, chronic renal insufficiency, myocardial infarction, heart failure, peripheral artery disease, chronic obstructive pulmonary disease (P>0.05).

**Conclusion:** The epidemiological status of hyperlipidemia in northern Henan Province had the specificity, the incident of hyperlipidemia was correlated to gender, ethnicity, age, marital status, education level, hypertension, and diabetes mellitus, those results may provide basis for the prevention and treatment for hyperlipidemia in northern Henan Province.

## Introduction

Hyperlipidemia is a kind of disease that endangers human health caused by high levels of blood lipids (including triglyceride, total cholesterol, and low-density lipoprotein cholesterol increased, and high-density lipoprotein cholesterol decreased in plasma) in the human body, according to ‘the Prevention and Treatment guidelines for Dyslipidemia in Chinese adults (revised in 2016)’, hyperlipidemia could be divided into four types, including hypercholesterolemia, hypertriglyceridemia, mixed hyperlipidemia and low high-density lipoprotein, the clinical manifestations of hyperlipidemia include xanthoma caused by lipid deposition in the dermis and arteriosclerosis caused by lipid deposition in the vascular endothelium [1]. In the normal human body, most patients do not have obvious symptoms or abnormal signs until blood biochemical tests are conducted to detect an increase in plasma lipoprotein levels, which undoubtedly increase difficulty to the prevention and treatment of hyperlipidemia.

It is reported that the average serum level of TC, LDL-C, and TG of adults in 2018 in China were significantly increased compared with the data obtained from the national survey in 2015; moreover, the data demonstrated that the average levels of TC and non HDL-C in Chinese adults are approaching or exceeding the average level of some western countries [2, 3], and it is predicted that an increase in serum cholesterol levels in the Chinese population is expected to lead to an increase of approximately 9.2 million cardiovascular events in China from 2010 to 2030 [4], those evidences demonstrated that hyperlipidemia is becoming a chronic noninfectious disease diseases that threaten human life and health, to understand the epidemiological status and risk factors of hyperlipidemia is necessary for prevention and treatment of hyperlipidemia. For this purpose, we collected and analyzed the clinical data in Xinxiang Central Hospital from 2019-2021, hope to provide some useful information for prevention and treatment of hyperlipidemia in north Henan Province.

## Methods

A total of 670 clinical data in Xinxiang Central Hospital from 2019-2021 were collected, and those data were classified and analyzed according to the basic information of investigation subjects including gender, ethnicity, age, smoking, marital status, occupation, education level and risk factors including hyperlipidemia, diabetes mellitus, ischemic stroke, coronary heart disease, angina pectoris, myocardial infarction, heart failure, peripheral artery disease, chronic renal insufficiency, gastrointestinal ulcers, chronic obstructive pulmonary disease, cerebral hemorrhage.

## Results

### 1. The correlation between general information of investigation subjects and hyperlipidemia in North Henan Province

The results demonstrated that there were 530 non-hyperlipidemia people and 130 hyperlipidemia people, accounting for 79.10% and 19.40% in the total investigation population, respectively; there were 96 male hyperlipidemia people and 44 female hyperlipidemia people, accounting for 68.57% and 31.43% in the total hyperlipidemia people, respectively; there were 664 Han ethnicity people and 6 Hui ethnicity people, accounting for 99.10% and 0.90% in the total investigation population, respectively; the age range of investigation population was 20-97 year old, and the number of people aged between 61-80 year old was highest (340/670, 50.75%); there were 390 people with a history of smoking, accounting for 58.21% in the total investigation population, 346 people were never smoking, accounting for 51.64% in the total investigation population; in this investigation, the proportion of peasant farmers was the highest (353/670, 52.69%), followed by retire population (164/670, 24.48%); in terms of education level, the proportion of people graduating from middle school was the highest (338/670, 50.45%), followed by those graduating from primary school (249/670, 37.16%); the statistical results found that the incident of hyperlipidemia were correlated to gender (P<0.01), ethnicity (P<0.01), age (P<0.05), marital status (P<0.05), and education level (P<0.001) (Table 1).

**Table 1.** The correlation between general information of investigation subjects and hyperlipidemia in North Henan Province.

| General information |  | Non-hyperlipidemia ( n=530) |  | Hyperlipidemia (n=140) |  | Significance /P value |
| --- | --- | --- | --- | --- | --- | --- |
|  |  | n | % | n | % |  |
| Gender | Male | 425 | 80.19% | 96 | 68.57% | 0.0043** |
|  | Female | 105 | 19.81% | 44 | 31.43% |  |
| Ethnicity | Han | 524 | 98.87% | 140 | 100% | 0.0043** |
|  | Hui | 6 | 1.13% | 0 | 0.00% | 0.3528 |
| Age | 20-40 | 21 | 3.96% | 2 | 1.43% | 0.3528 |
|  | 41-60 | 173 | 37.27% | 47 | 28.53% | 0.1933 |
|  | 61-80 | 268 | 50.57% | 72 | 51.43% | 0.0171* |
|  | ≥81 | 68 | 12.83% | 19 | 13.57% | 0.9243 |
| Smoking | Currently smoking | 220 | 41.51% | 48 | 34.29% | 0.7792 |
|  | Quit smoking ≥ 1 years | 33 | 6.23% | 8 | 5.71% | 0.1455 |
|  | Quit smoking ≥ 1 months | 15 | 2.83% | 0 | 0.00% | >0.9999 |
|  | Never smoking | 262 | 49.43% | 84 | 60.00% | 0.05* |
| Marital status | Married (Remarried) | 503 | 94.91% | 135 | 96.43% | 0.0288* |
|  | Single | 3 | 0.57% | 0 | 0.00% | 0.6553 |
|  | Bereave | 19 | 3.58% | 5 | 3.57% | >0.9999 |
|  | Divorce | 5 | 0.94% | 0 | 0.00% | >0.9999 |
| Occupation | Peasant farmer | 273 | 51.51% | 80 | 57.14% | 0.5984 |
|  | Worker | 83 | 15.66% | 19 | 13.57% | 0.2541 |
|  | Civil servants/<br>Government officials | 6 | 1.13% | 2 | 1.43% | 0.5984 |
|  | Retirees | 129 | 24.34% | 35 | 25.00% | 0.6753 |
|  | Professional technical personnel | 8 | 1.51% | 0 | 0.00% | 0.9121 |
|  | Individual business owners | 6 | 1.13% | 1 | 0.71% | >0.9999 |
| Education level | Illiteracy | 31 | 5.85% | 2 | 1.43% | 0.0282* |
|  | Primary school | 179 | 33.77% | 70 | 50.00% | 0.0006*** |
|  | Middle School/Vocational School | 273 | 51.51% | 65 | 46.43% | 0.2971 |
|  | College/University | 46 | 8.68% | 2 | 1.43% | 0.0014** |
|  | Postgraduate | 1 | 0.19% | 1 | 0.71% | 0.3745 |

\*P<0.05, \*\*P<0.01, \*\*\*P<0.0001.

### 2. The correlation between risk factors and hyperlipidemia in north Henan Province

And then we analyzed the correlation between risk factors and hyperlipidemia, found that the incident of hyperlipidemia was correlated to hypertension and diabetes mellitus, the P value were 0.0101 and 0.0001, respectively, had no relation to cerebral hemorrhage, angina pectoris, ischemic stroke, gastrointestinal ulcers, coronary heart disease, chronic renal insufficiency, myocardial infarction, heart failure, peripheral artery disease, chronic obstructive pulmonary disease (P>0.05) (Table 2).

**Table 2.** The correlation between risk factors and hyperlipidemia in north Henan Province.

| Risk factors | Non-hyperlipidemia ( n=530) |  | Hyperlipidemia (n=140) |  | Significance/<br>P value |
| --- | --- | --- | --- | --- | --- |
|  | n | % | n | % |  |
| Hypertension | 255 | 48.11% | 85 | 60.71% | 0.0101* |
| Diabetes mellitus | 85 | 16.04% | 47 | 33.57% | <0.0001*** |
| Ischemic Stroke | 73 | 13.77% | 28 | 20.00% | 0.0834 |
| Coronary heart disease | 4 | 0.75% | 1 | 0.71% | >0.9999 |
| Angina pectoris | 89 | 16.79% | 15 | 10.71% | 0.088 |
| Myocardial infarction | 39 | 7.36% | 6 | 4.29% | 0.2548 |
| Heart Failure | 8 | 1.51% | 1 | 0.71% | 0.6932 |
| Coronary intervention | 24 | 4.53% | 2 | 1.43% | 0.1362 |
| Peripheral Artery Disease | 1 | 0.19% | 1 | 0.71% | 0.3745 |
| Chronic renal insufficiency | 3 | 0.57% | 1 | 0.71% | >0.9999 |
| Gastrointestinal ulcers | 15 | 2.83% | 1 | 0.71% | 0.2146 |
| Chronic Obstructive<br>Pulmonary Disease | 7 | 1.32% | 1 | 0.71% | >0.9999 |
| Cerebral hemorrhage | 6 | 1.13% | 1 | 0.71% | >0.9999 |

\*P<0.05, \*\*P<0.01, \*\*\*P<0.0001.

## Discussion

Hyperlipidemia, also known as dyslipidemia, is usually associated with elevated triglycerides and total cholesterol in plasma, is developing into one of the main diseases that affect people’s daily health. Hyperlipidemia can be divided into primary hyperlipidemia and secondary hyperlipidemia according to pathogenesis, it is reported that hyperlipidemia could occur at different ages and genders, the incidence rate of hyperlipidemia was increased with the age, and the highest incidence occurs between 50-69 years old; many factors including gender [5, 6], alcohol consumption [7, 8], smoking [9], diet [10], physical activity [11], and emotional activity [12] could affect the occurrence of hyperlipidemia. In our data, we also found that the incident of hyperlipidemia was correlated to gender (P<0.01), ethnicity (P<0.01), and age (P<0.05) in north Henan province, these results are consistent with the previously reported results, taking regular exercise and dietary therapy may effectively decrease or alleviate the occurrence of hyperlipidemia; furthermore, our data also demonstrated that the marital status (P<0.05) and education level (P<0.001) also have the higher correlation to the incident of hyperlipidemia, there is a significant difference in the incidence of hyperlipidemia between married populations and unmarried populations, between high educated populations and low educated populations, the reason for this result may because married populations and unmarried populations, high educated populations and low educated populations have the different economic situation and dietary habits, but the further research is needed.

Hyperlipidemia is closely related to hypertension and diabetes mellitus, in many cases, hypertension and diabetes mellitus also could result in hyperlipidemia, it is reported that hyperlipidemia is also the main risk factor of hypertension and diabetes mellitus [13, 14]. In our data, we found that the incident of hyperlipidemia was highly correlated to hypertension (P=0.0101) and diabetes mellitus (P=0.0001) in north Henan Province, these evidences demonstrated we should strengthen the medical care and management of people with hypertension and diabetes mellitus, simultaneously combined with taking regular exercise and dietary therapy to decrease the incidence of hyperlipidemia.

## Conclusion

The epidemiological status of hyperlipidemia in northern Henan Province had the specificity, the incident of hyperlipidemia was correlated to gender, ethnicity, age, marital status, education level, hypertension, and diabetes mellitus, those results may provide treatment or prevention reference guide for hyperlipidemia in northern Henan Province.

## Data Availability

All data produced in the present study are available upon reasonable request to the authors

## Acknowledgements

None.

